# Development of a quantum-dot lateral flow immunoassay strip based portable fluorescence smart-phone system for ultrasensitive detection of IgM/IgG to SARS-CoV-2

**DOI:** 10.1101/2020.07.21.20159392

**Authors:** Bochao Liu, Jinfeng Li, Xi Tang, Ze Wu, Jinhui Lu, Chaolan Liang, Shuiping Hou, Ling Zhang, Tingting Li, Wei Zhao, Yongshui Fu, Yuebin Ke, Chengyao Li

**Author notes:** Corresponding author: Chengyao Li or Yuebin Ke, School of Laboratory Medicine and Biotechnology, Southern Medical University. No. 1838 North Guangzhou Avenue, Guangzhou 510515, China., (Li C) or (Ke Y). These authors contributed equally to this work (Bochao Liu, Jinfeng Li, Xi Tang).

## Abstract

**Background:** Since December 2019, the outbreak of coronavirus disease (COVID-19) has been occurred by novel coronavirus (SARS-CoV-2). The rapid and sensitive immunoassays are urgently demanded for detecting specific antibodies as assistant diagnosis for primary screening of asymptomatic individuals, close contacts, suspected or recovered patients of COIVD-19 during the pandemic period.

**Methods:** The recombinant receptor binding domain of SARS-CoV-2 spike protein (S-RBD) was used as the antigen to detect specific IgM and the mixture of recombinant nucleocapsid phosphoprotein (NP) and S-RBD were used to detect specific IgG by the newly designed quantum-dot lateral flow immunoassay strip (QD-LFIA), respectively.

**Results:** A rapid and sensitive QD-LFIA based portable fluorescence smart-phone system was developed for detecting specific IgM/IgG to SARS-CoV-2 from 100 serum samples of COVID-19 patients and 450 plasma samples from healthy blood donors. Among 100 COVID-19 patients diagnosed with NAT previously, 3 were severe, 35 mild and 62 recovered cases. By using QD-LFIA, 78 (78%) and 99 (99%) samples from 100 COVID-19 patients’ serum were detected positive for anti-SARS-CoV-2 IgM or IgG, respectively, but only one sample (0.22%) was cross-reactive with S-RBD from 450 healthy blood donor plasmas that were collected from different areas of China.

**Conclusion:** An ultrasensitive and specific QD-LFIA based portable fluorescence smart-phone system was developed fo r detection of specific IgM and IgG to SARS-CoV-2 infection, which could be used for investigating the prevalence or assistant diagnosis of COVID-19 in humans.

## INTRODUCTION

In December 2019, a coronavirus pneumonia (COVID-19) was firstly discovered in Wuhan, China and then largely reported within a few months in the rest of world, which was caused by the novel coronavirus designated as SARS-CoV-2 [1,2]. The small part of patients rapidly developed acute complications such as acute respiratory distress syndrome (ARDS) and acute respiratory failure [3]. The main routes of SARS-CoV-2 transmission in humans were respiratory droplets and virus exposure between individuals. By the middle of July 2020, more than 13 million people were diagnosed as COIVD-19 by viral nucleic acid test (NAT) and over 500 thousand deaths were announced globally, among them more than 80 thousands cases and 4000 deaths were reported in China. SARS-CoV-2 has been found more infectious than SARS coronavirus, especially for the middle-aged and elderly people [4]. Due to the existence of close contacts, suspected cases and asymptomatic patients or resolvers of COIVD-19 can be discovered by detecting the antibodies specific to SARS-CoV-2.

The SARS-CoV-2 is a single strand sense RNA virus and belongs to β genus of coronavirus family [5–7], and has nearly 30,000 bases of nucleotides in length [8]. Based on the known genome sequence (GenBank MN908947), testing of SARS-CoV-2 RNA was commonly used for detection of virus. Due to the stability of reagents and primers, some nucleic acid tests were not sensitive enough to detect SARS-CoV-2 from nasal or pharynx swabs [9–12]. Many patients had to be tested repeatedly before they were finally diagnosed [13,14]. However, testing of specific antibodies to SARS-CoV-2 can be a helpfully assistant method.

Serum IgM or IgG specific to SARS-CoV-2 might co-exist with viral RNA, which could be used for preliminary screening and auxiliary diagnosis of COVID-19 [15]. A previous study reported that less than 40% of antibodies were presence within 1-week among patients since onset, and rapidly increased to 100% (either IgM or IgG), 94.3% (IgM) and 79.8% (IgG) since day-15 after onset, while IgM declined to the cutoff level in 39 days but IgG persisted in the high level [16,17]. Several antibody assays have been developed for detection of SARS-CoV-2 infection [18–21]. However, the more specific, sensitive, rapid and simple tests are still required strongly. In this study, we designed a quantum-dot lateral flow immunoassay strip (QD-LFIA) based potable fluorescence smart-phone system for detection of specific IgM/IgG to SARS-CoV-2 in human serum samples. This assay has two innovative techniques: (1) a new format assay includes a portable fluorescence detection platform which can be operated in biological safety hoods or cabinets for avoiding possible aerosol contamination, and in wireless connecting smart-phone for processing data efficiently; (2) the testing can be carried out within 15 min. This novel assay is ultrasensitive, cost-effective, rapid and simple, which can be used for on-site detection of serum IgM/IgG specific to SARS-CoV-2 infection in humans.

## MATERIALS AND METHODS

### Blood specimen

The numbers of 100 serum samples of COVID-19 patients were provided by the Shenzhen Center for Disease Control and Prevention (CDC), which were confirmed positive for SARS-CoV-2 RNA by real-time RT-PCR (RT-qPCR). A total of 450 plasma samples of healthy blood donors were obtained from blood centers of Shenzhen and Guangzhou (south), Harbin (northeast), Chengdu (southwest) and Xi’an (northwest), which were collected during the period of August to October of 2019 before the COVID-19 outbreak in Wuhan (middle), China. These blood donor samples were used as negative control. This study was approved by the Medical Ethics Committees of Shenzhen CDC and Southern Medical University, and followed the ethical guidelines of the 1975 Declaration of Helsinki.

### Reagents and equipment

Bovine serum albumin (BSA) and goat anti-mouse polyclonal immunoglobulin G (G-mIgG) were purchased commercially (Sigma-Aldrich, St. Louis, USA). Mouse anti-human IgG (M-hIgG) and IgM (M-hIgM) were purchased from Beijing Bersee Science and Technology Co. Ltd (Beijing, China). Recombinant spike protein receptor binding domain fused with mouse IgG-Fc fragment (S-RBD) and nucleocapsid phosphoprotein (NP) of SARS-CoV-2 were purchased from Bioeast Biotech Co. Ltd (Hangzhou, China) and S-RBD was produced from HEK293 cells and NP was produced in *E*. *coli*, respectively. Polystyrene-coated quantum dot nanoparticles (QDs) with 160 nm diameter were purchased from Huge Biotech Ltd (Shanghai, China). Nitrocellulose (NC) membranes, glass fibers and absorbent pads were purchased from Millipore Corporation (Bedford, MA, USA). A 3D-printed portable fluorescence strip reader was designed by this study. The colloid gold lateral flow immunoassay strip (CG-LFIA; Lvshiyuan Biotechnology Co. Ltd., Shenzhen, China) was used as comparative assay for detection of antibodies to SARS-CoV-2.

### Preparation of QD-LFIA strips

QD-LFIA strip was composed of a 25 mm sample pad, 20 mm NC membrane and 15 mm absorbent pad, and all of those were pasted on an adhesive backing card. The conjugate pad was saturated with treatment buffer (10mM PBS, pH 7.4 containing 4% sucrose, 0.5% Tween-20, 0.5% PVP and 1% BSA) for 1 h and dried at 37 °C for 24h.

An aliquot of QDs (25μL) were conjugated with 10 μg M-hIgG or S-RBD by covalent attachment, respectively. The M-hIgG@QD and S-RBD@QD conjugates were mixed, and then dispensed onto the conjugate pad at 2μL/cm by a Biolet XYZ-3060 Quantidispenser, followed by drying at 37°C before storing in a dry cabinet at room temperature for ready use. The G-mIgG (0.5μg/cm), mixture of M-hIgG and NP (0.5μg each protein/cm) or M-hIgM (1μg/cm) were spotted on the NC membrane (from absorbent pad to sample pad direction) to generate the C, T2 and T1 lines, respectively (Fig. 1A). The coated NC was dried at 65 °C for 30min. The hemofiltration sample pad was pre-treated with the blocking buffer (10mM PBS, pH 7.4 containing 2% sucrose, 0.4% Tween-20, 0.5% S9 and 10% casein) and dried at 37°C for overnight.

**Fig 1.**
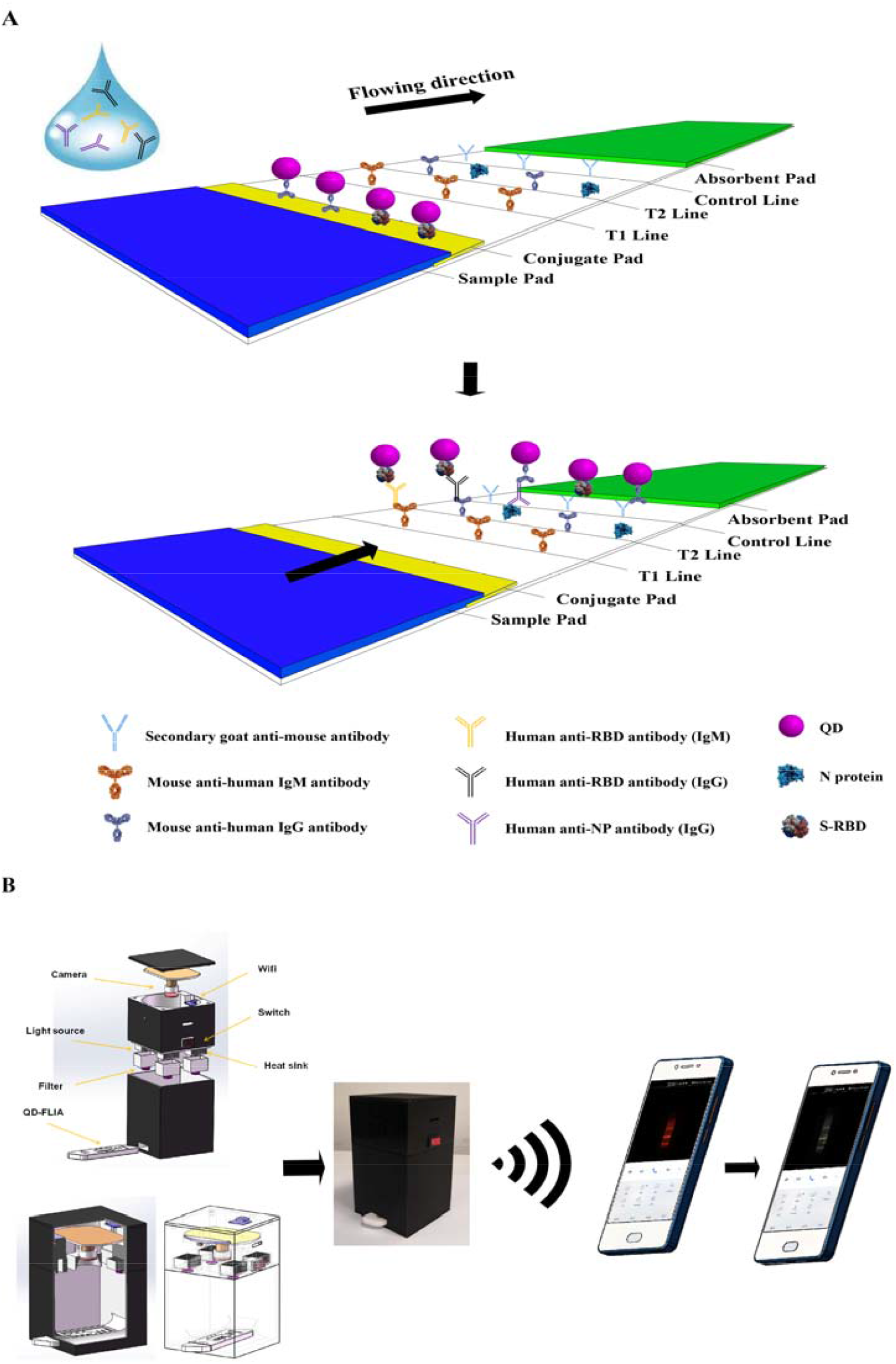
The schematic diagram of QD-LFIA (A) based portable fluorescence smart-phone system (B) for detection of specific IgM/IgG to SARS-CoV-2 infection in humans.

**Fig 2.**
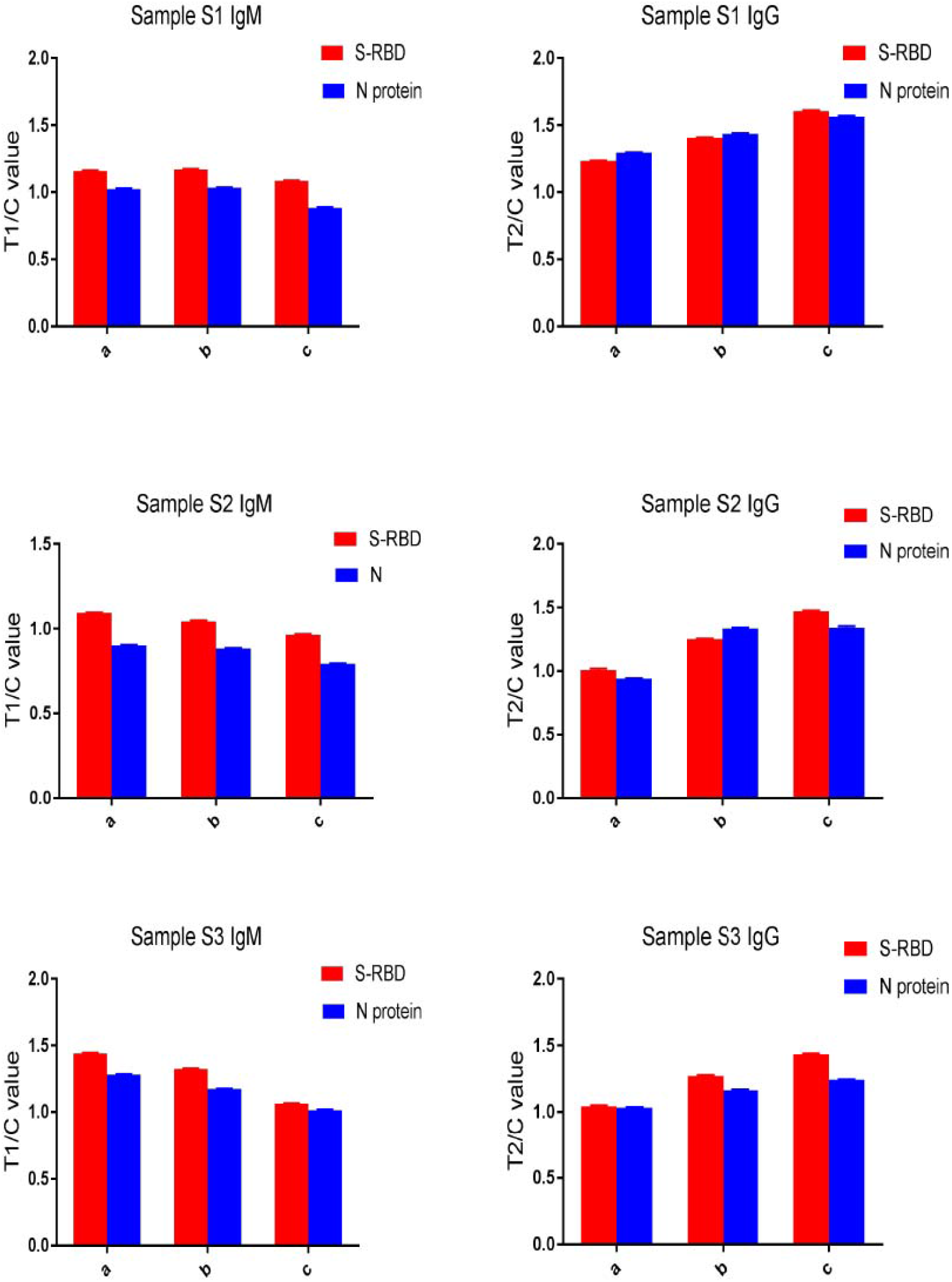
T/C values of QD-LFIA for detection of serum IgM or IgG reactive to S-RBD and NP of SARS-CoV-2 from three severe COVID-19 patients (samples S1-S3). S-RBD and NP were used as detection antigens in the assay.

QD-LFIA testing was performed by mixing 1μL of serum sample with 120μL sample buffer (10mM PBS, pH 7.4 containing 0.05% Tween-20 and 1% BSA). Then the reactive sample solution was taken onto sample pad of the testing strip for immunochromatography. Fluorescence signals on T1/T2 and C lines of strip were measured in 10 min by a portable fluorescence strip reader and were uploaded to the smart-phone via wifi for processing the data.

### Statistical analysis

All experiments were performed at least three times independently. The data were analyzed using the statistical package SPSS v. 16.0. The results were presented as the mean±SD. Difference between groups was analyzed by using Student’s t-test, and *P*-value < 0.05 was considered statistically significant.

## RESULTS

### Invention of QD-LFIA based portable fluorescence smart-phone system

In order to report the testing results effectively, a portable fluorescence smart-phone system was invented for QD-LFIA (Fig.1). This system included a QD-LFIA strip with C and T1/T2 lines, a portable fluorescence strip reader with laser, real-time camera and wifi, and a mobile phone with signal receiver and data processor. When a reactive strip was inserted into the fluorescence reader, the fluorescent images on T1/T2/C lines were taken and transited to a mobile smart-phone via wifi. The fluorescence lines were selected and the fluorescent intensity of T1, T2 or C lines was converted to the peak area. The value of IgG/control (T2/C) or IgM/control (T1/C) was calculated, and then the reaction of sample was reported for negativity or positivity after comparing with the cut-off value by artificial intelligence (AI) processor.

### Detection of COVID-19 IgM/IgG by QD-LFIA

Fifty healthy blood donor plasma samples (provided by Guangzhou blood center) were tested negative for IgM/IgG to SARS-CoV-2 by ELISA. By using these negative control samples, the appropriate cut-off values of IgM/IgG in QD-FLIA system were defined as 0.0964 or 0.0952 (mean plus 3 SD), respectively. In comparison with CG-LFIA, 100 serum samples of COVID-19 patients and 450 plasma samples of healthy blood donors were detected by QD-LFIA for specific IgM/IgG to S-RBD or NP mixture, respectively. Among 100 COVID-19 cases diagnosed previously, 3 were severe, 35 mild and 62 recovered patients. Each of three severe patients had 3 follow-up samples (S1-S3), containing the high level of both IgM and IgG responses to either S-RBD or NP in QD-LFIA (Fig.2). Among 35 mild patients, 32 positive IgM to S-RBD and 22 positive IgM to NP were detected by QD-LFIA, in which S-RBD presented higher IgM response than NP in QD-LFIA excepting for three cases (Fig. 3A and 3B, *P*<0.05). For detection of specific IgG among 35 mild patients, 30 cases were tested positive by S-RBD or NP based QD-LFIA, respectively (Fig. 3C and 3D), while each antigen based QD-LFIA had 5 positive samples uniquely, but the combined S-RBD and NP antigens were all positive for 35 samples. Among 62 recovered patients, specific IgM was detected in 43 cases by S-RBD-based QD-LFIA and 9 cases by NP-based QD-LFIA, overall were 43 cases by two antigens but no complementary (Fig. 4A and 4B). For IgG detection, 61/62 recovered patients were detected positive by the combined S-RBD and NP antigens in QD-LFIA, while a sample was negative by QD-LFIA (Fig.4C and 4D). The reactivity of IgG to S-RBD or NP was similar among recovered patients, in which 10 samples were higher reactive to S-RBD but 8 to NP, respectively.

**Fig 3.**
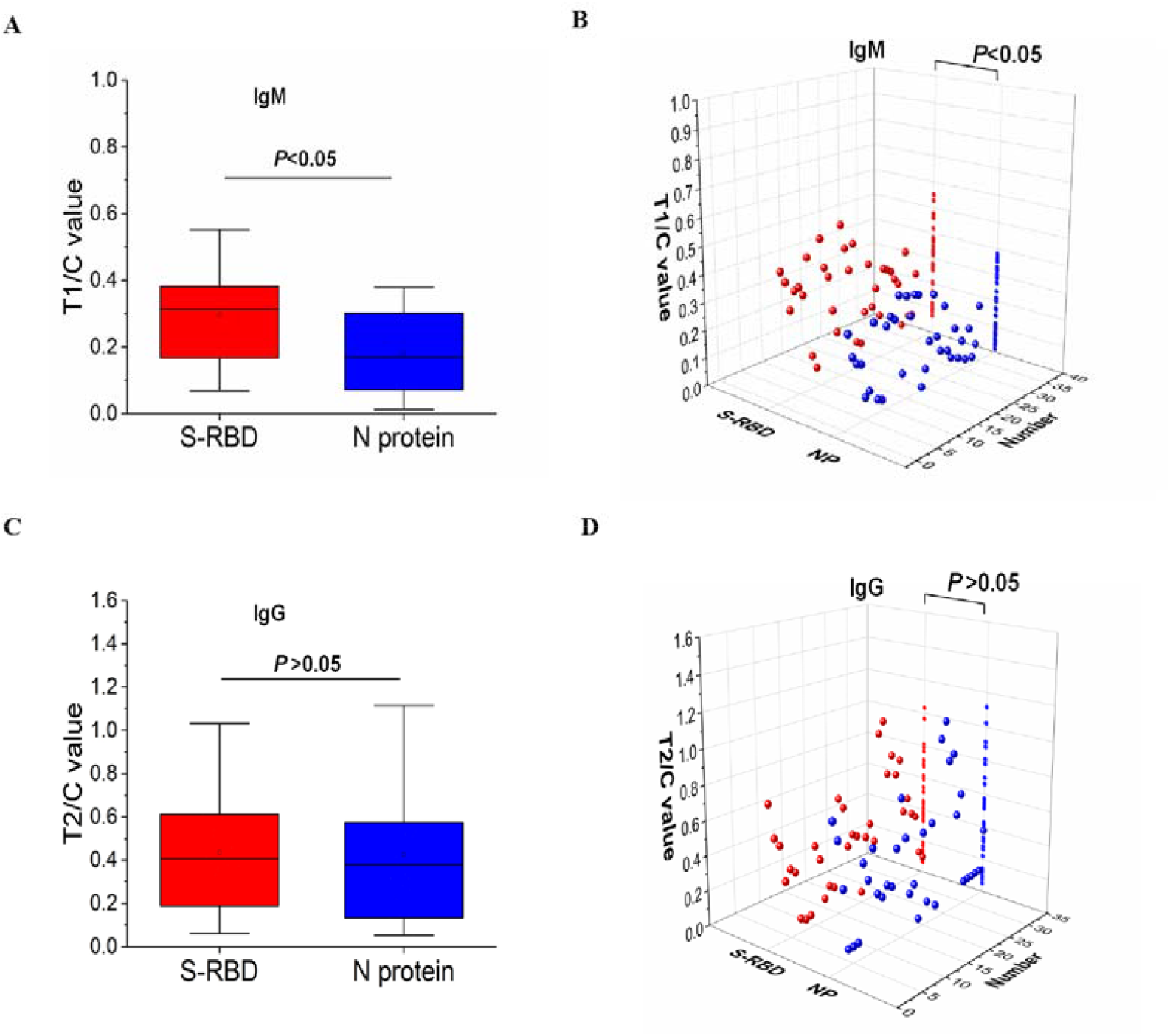
T/C values of QD-LFIA for detection of serum IgM (A, B) or IgG (C, D) specific to SARS-CoV-2 from 35 mild COVID-19 patients, respectively. S-RBD and NP were used as detection antigens in QD-LFIA. * *P* < 0.05.

**Fig 4.**
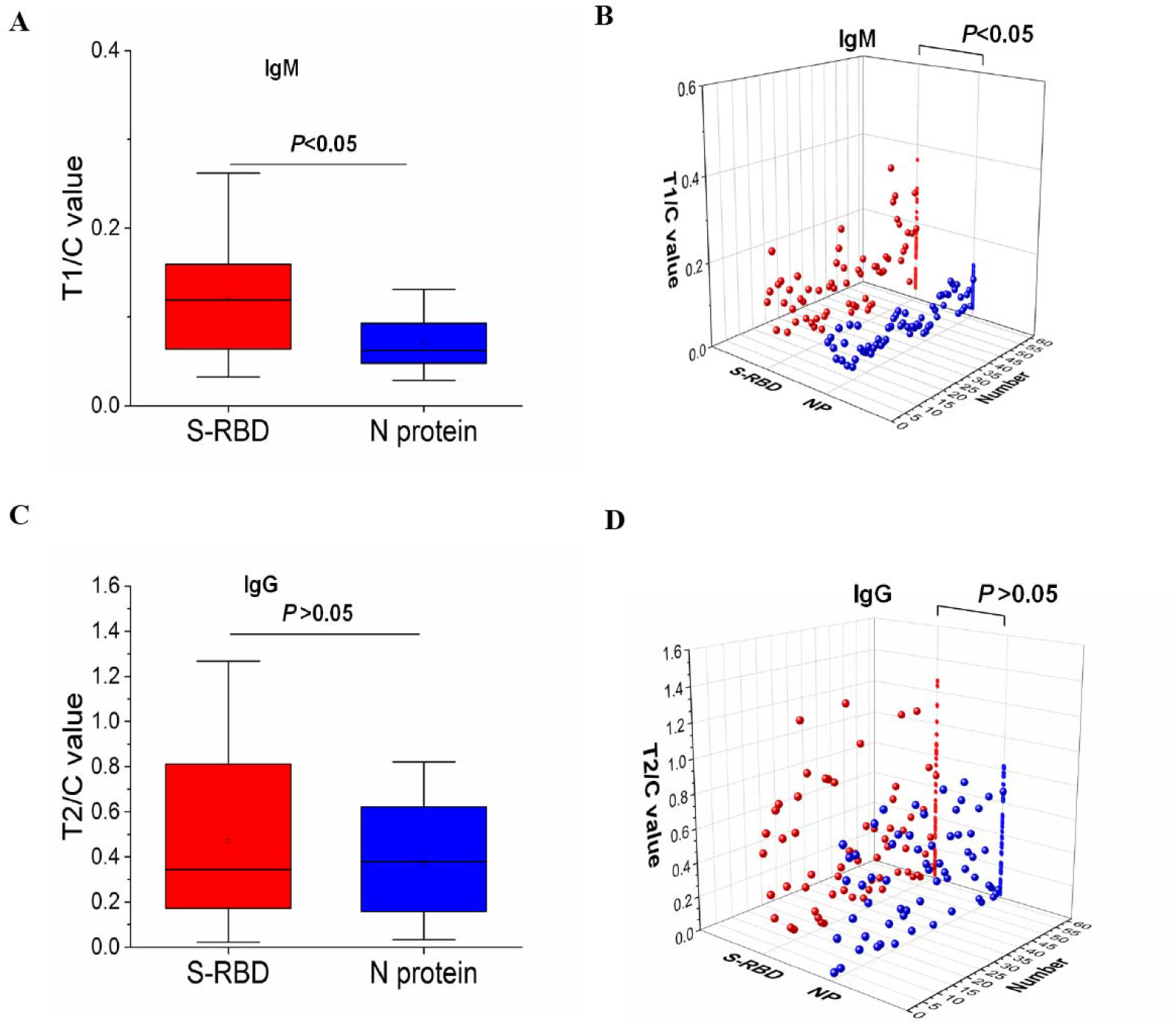
T/C values of QD-LFIA for detection of serum IgM (A, B) or IgG (C, D) reactive to SARS-CoV-2 from 62 recovered COVID-19 patients using S-RBD or NP as detection antigens, respectively. * *P* < 0.05.

**Fig 5.**
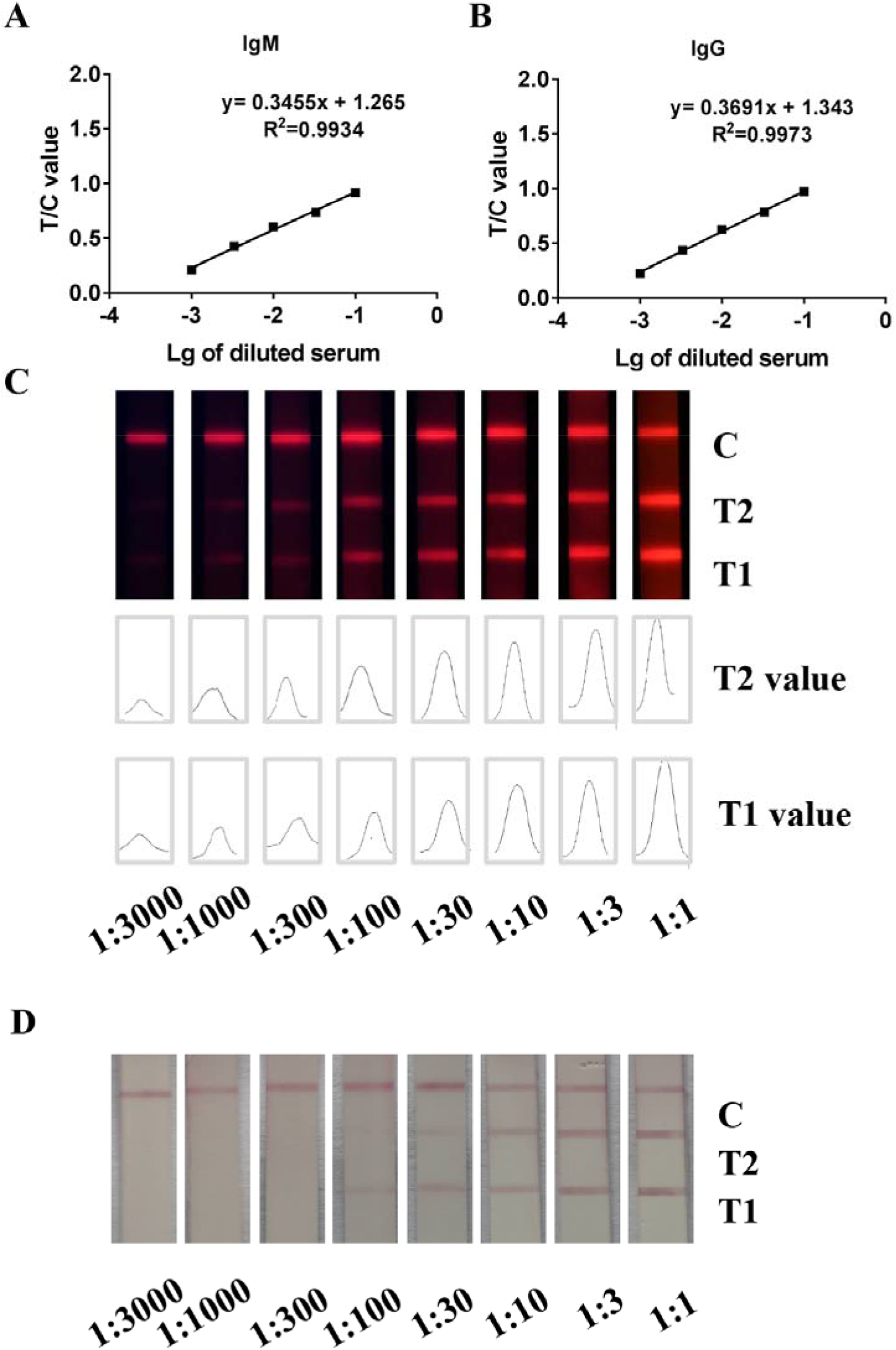
QD-LFIA linear regression of quantitative testing of IgM/IgG to SARS-CoV-2 in blood samples from COVID-19 patients. (A, B) The linear regression curves for IgM and IgG in serum sample. (C) The fluorescent images of reactive C, T2and T1 lines and the converted areas of T1/T2 fluorescence density in QD-LFIA. (D) The images of reactive C, T2 and T1 lines in CG-LFIA.

In addition to 50 Guangzhou blood donors, 400 plasma samples of healthy blood donors from South, Southwest, Northeast and Northwest of China were tested for reactive IgM/IgG to S-RBD and NP antigens by QD-LFIA, respectively. All samples were negative to S-RBD and NP based QD-LFIA excepting for that a sample from Chengdu (Southwest of China) had IgG reactive to S-RBD in QD-LFIA.

### Sensitivity and specificity of QD-LFIA

According to above results obtained by two kinds of S-RBD and NP antigens, S-RBD presented better sensitivity than NP to IgM in QD-LFIA, while S-RBD and NP had mutual complementary reactivity for detection of specific IgG in QD-LFIA. Compared with conventional CG-LFIA, one serum sample with serial tenfold dilutions were measured for reacting with IgM or IgG by QD-LFIA, which showed that QD-LFIA detected IgM/IgG in 1:1000 diluted sample (Fig.5), which was 10-100 times sensitive than CG-LFIA. Among 100 COVID-19 cases diagnosed previously by NAT at hospitals, the detection rate for reactive IgM was 78% (78/100) by S-RBD based QD-LFIA, and for reactive IgG was 99% (99/100) by the combined S-RBD and NP based QD-LFIA (Table 1). From comparison with control assay, CG-LFIA only had 32% of IgM and 71% of IgG positivity (Table 1).

**Table 1.** Sensitivity and specificity of QD-LFIA for testing of anti-SARS-CoV-2 IgM and IgG

| Population | Sample<br>(Nb*) | QD-LFIA |  | CG-LFIA |  |
| --- | --- | --- | --- | --- | --- |
|  |  | (Nb/%) |  | (Nb/%) |  |
| COVID-19 |  | IgM | IgG | IgM | IgG |
| Severe | 3 | 3 (100%) | 3 (100%) | 3 (100%) | 3 (100%) |
| Mild | 35 | 32 (94.4) | 35 (100%) | 23(65.7%) | 26(74.3%) |
| Recovered | 62 | 43(69.4%) | 61(98.4%) | 6(9.7%) | 42(67.7%) |
| Overall | 100 | 78 (78%) | 99 (99%) | 32 (32%) | 71(71%) |
| Blood donor | 450 | 0 (0) | 1 (0.22%) | 0 (0) | 1 (0.22%) |

\* Number.

For identifying the specificity of QD-LFIA, none (0/450) of cross-reactivity was found IgM positive and only one sample (1/450) was detected IgG positive from 450 healthy blood donors, which suggested that the specificity of QD-LFIA was 100% for IgM and 99.8% for IgG detection, which were identical with conventional CG-LFIA (Table 1).

## DISCUSSION

A brand new coronavirus from a patient’s pharyngeal swab sample was discovered by the Chinese Center for Disease Control and Prevention (CDC) on January 7, 2020, which was temporarily named 2019-nCoV, and subsequently designated as SARS-CoV-2 by the World Health Organization (WHO) [22]. SARS-CoV-2 has caused a global pandemic with novel coronavirus pneumonia disease (COVID-19). More than 5 millions of people are diagnosed with COVID-19 and over 300 thousands of deaths are reported so far. This pandemic has made many communities or cities closed, which caused huge economic losses worldwide. Currently, the re-opening or re-starting of social activities are initiating in Asian and European countries where the SARS-CoV-2 infection has been contained. Along with nucleic acid testing of SARS-CoV-2, the antibody tests are largely needed as well. The development of rapid and accurate antibody detection methods is highly welcomed. Detection of specific IgM and IgG to SARS-CoV-2 can reflect the status of virus infection among different populations, and also can provide serological evidence for clinical diagnosis of COVID-19 patients.

The colloid gold lateral flow immunoassay strip (CG-LFIA) is a commonly used rapid immunochromatography assay for antibody detection. It normally takes 10-15 minutes to report result without requiring of special equipment. However, CG-LFIA has the lower sensitivity and the testing is judged by eyes, which may give false result by human errors. In this study, we developed a quantum-dot based LFIA (QD-LFIA) and combined with a portable fluorescence real-time camera reader, which increased its sensitivity in 10-100 times than conventional CG-LFIA. For detection rates of specific IgM/IgG from 100 COVID-19 patients diagnosed by NAT previously, QD-LFIA identified 78% of IgM and 99% of IgG reactive samples, while CG-LFIA had only 32% of IgM and 71% of IgG reactive to SARS-CoV-2 antigens in patients’ sera, respectively (*P*<0.05). In view of biosafety, the testing and reading of QD-LFIA strip could be performed in the biosafety hood, while the data could be transmitted via wifi and received outside the closed experiment cabinet by a mobile smart-phone within the laboratory. It’s only takes 15min to obtain the results safely.

The S-RBD or NP of SAR-CoV-2 in single and combined forms were examined as antigens to detect specific IgM or IgG of blood samples from COVID-19 patients in this study (Fig. 1-4). According to the reactive rate, S-RBD was finally determined to detect IgM and S-RBD and NP mixture was used to detect IgG in QD-LFIA. No cross-reaction of these S-RBD and NP antigens was found with IgM but 1 (0.22%) case with IgG from 450 healthy blood donors, which suggesting that the specificity of QD-LFIA was 100% for IgM and 99.78 % for IgG for detection of antibodies from COVID-19 patients. Also, this cross-reactive IgG sample from a 45 years old woman (Chengdu, Southwest) was tested positive by the control assay CG-LFIA, which was considered as non-specific to S-RBD of SARS-CoV-2, indicating the donor might be infected previously by a general flu coronavirus such as human coronavirus 229E (HuCoV-229E), NL63 (*Alphacoronavirus*), HKU1 or OC43 (*Betacoronavirus*) (23), but not identified yet.

In conclusion, a QD-LFIA based portable fluorescence smart-phone system was invented for detecting specific IgM and IgG to SARS-CoV-2 infection, which could be applied for investigating the epidemiology or assistant diagnosis of COVID-19 in epidemic areas of world.

## Data Availability

All data associated with this study are available in the main text, and available in the Department of Transfusion Medicine, Southern Medical University, Guangzhou, China

## Acknowledgments

The author thanks for Guangzhou, Shenzhen, Chengdu, Haerbing and Xian blood centers for their providing the healthy blood donor samples.

This study has partly received the financial and technical supports from Guangzhou Bai Rui Kang or Breakthrough (BRK) Bioscience and Technology Limited Company, Guangzhou, China.

## Author contribution

C.L., Y.K. and B.L. designed study; B.L., J.L., X.T.,Z.W. and S.H. performed experiments; J.L., B.L., C.L. and T.L. analyzed data; Y. F. provided materials and B.L., L.Z., C.L. and Y.K. wrote the paper.

